# Smartphone Missingness as a Depression Biomarker: A Baseline-Controlled Re-analysis of StudentLife

**DOI:** 10.64898/2026.04.30.26351977

**Authors:** Ceyhun Olcan

**Affiliations:** Center for Technology and Behavioral Health, Geisel School of Medicine at Dartmouth, Lebanon, NH 03766, USA

**Keywords:** digital phenotyping, passive sensing, smartphone sensing, depression, PHQ-9, missing data, baseline-controlled prediction, cross-validation, ecological momentary assessment, StudentLife

## Abstract

**Background:** Whether gaps in smartphone passive-sensing data carry psychological signal — beyond what a baseline self-report already provides — is a recurring question in digital phenotyping that has rarely been tested with multiplicity control and cross-validation on the same cohort.

**Objective:** To test whether participant-level missingness in passive sensing and ecological momentary assessment (EMA) is an incremental predictor of depression beyond a single baseline self-report, in the canonical StudentLife cohort.

**Methods:** Exploratory re-analysis of the publicly available StudentLife dataset. The original 2014 publication reports *n* = 48 enrolled undergraduates; the public archive contains 59 distinct sensor-instrumented UIDs, but the additional 11 are sparse-data or PHQ-9-incomplete records and contribute nothing to the between-person analyses, which use only the 38 participants with paired pre-term and post-term Patient Health Questionnaire-9 (PHQ-9). We computed 89 participant-level missingness features from nine continuous sensor streams, five phone-activity logs and 27 EMA prompts, and evaluated them under leave-one-out cross-validation with nested-CV-tuned hyperparameters, cluster-bootstrap confidence intervals, an omnibus joint *F*-test, and Benjamini–Hochberg multiplicity control.

**Results:** A pre-term PHQ-9 baseline alone explained 59% of out-of-sample variance in post-term scores (*n* = 32; 95% cluster-bootstrap CI [0.22, 0.81]). Tuned regularized linear models trained on missingness alone reached only the cohort-mean baseline; adding missingness to pre PHQ-9 did not improve performance. The omnibus joint *F*-test of all nine continuous-stream missingness rates against post-term PHQ-9, adjusted for pre-term PHQ-9, was non-significant (*F* (9, 27) = 0.43, *P* = 0.91). No individual feature survived multiplicity correction. A separate within-person day-level analysis (2,186 person-days) yielded a small valence-specific prospective effect (*r* = +0.082, 95% CI [+0.011, +0.162]) opposite in direction to the withdrawal hypothesis.

**Conclusion:** In this cohort, smartphone-data missingness did not add incremental predictive value beyond a single baseline PHQ-9. The analysis is exploratory and StudentLife-specific; it should not be read as evidence that missingness is never informative.

**Plain-language summary:** Many studies use the gaps in someone’s smartphone data — missing GPS readings, missed survey prompts, fewer phone interactions — as a possible warning sign of depression. This re-analysis tested that idea on a widely used public dataset from a class of 48 college students. After accounting for each student’s depression score at the start of the term, the gaps in their phone data added no useful information about their depression score at the end of the term. The result is specific to this dataset and does not mean that smartphone gaps are never informative, but it shows that such claims need careful baseline comparisons.

**Key Points:** *Question:* Is smartphone-data missingness an incremental depression biomarker beyond a single baseline self-report?

*Findings:* In an exploratory re-analysis of the StudentLife cohort (38 participants with paired PHQ-9 scores), missingness features did not improve prediction of post-term PHQ-9 beyond pre-term PHQ-9 under leave-one-out cross-validation, nested-CV-tuned models, an omnibus joint *F*-test, or multiplicity-controlled univariate screens.

*Meaning:* Smartphone-data missingness should not be interpreted as a psychological signal in absence of baseline-controlled, cross-validated, multiplicity-aware evidence. The result is specific to a high-functioning undergraduate cohort over a ten-week term and requires replication.

---

Smartphone-based passive sensing has become a dominant paradigm in digital mental health, with streams from accelerometers, GPS, microphones, Bluetooth, Wi-Fi, screen activity and phone-state events combined with self-report instruments to infer psychological state and, most prominently, depression ^1,2^. Standard practice is to treat the gaps in these streams as ignorable: drop affected participant-days, impute, or compute features only over observed segments.

Several lines of evidence have argued, more or less explicitly, that this ignorability is wrong in depression. Pratap and colleagues found that retention in remote digital health studies tracked clinical and demographic characteristics in ways that complicate naï ve imputation ^3^; Saeb et al. reported that mobile-phone-derived behavioural disengagement (e.g., reduced GPS-derived location entropy) correlated with depressive symptom severity ^4^; Wang et al. derived weekly behavioural features from StudentLife sensor data that tracked DSM-5-based depressive symptoms ^5^. Each of these claims rests, implicitly or explicitly, on the idea that withdrawal from device wear and from study participation is a downstream behavioural manifestation of the same depressive process that produces clinical symptoms. If correct, missingness is a candidate digital biomarker with a relatively direct mechanistic interpretation rather than a nuisance.

The missingness-as-signal hypothesis is attractive but is rarely tested directly with the level of statistical control normally required of a biomarker claim. A defensible test requires four ingredients that are frequently absent in the literature: (i) explicit comparison to a baseline self-report on the same outcome, so that any apparent gain from missingness can be attributed to genuine incremental information rather than to a baseline that the model has rediscovered through proxy features; (ii) cross-validated rather than in-sample predictive evaluation, so that reported performance estimates are not optimistically biased; (iii) multiplicity control across the (often very large) feature set, since most published missingness analyses screen tens of features against a single outcome; and (iv) an omnibus association test rather than reliance solely on per-feature univariate screens.

The StudentLife dataset ^6^, conducted at Dartmouth College in spring 2013, is a canonical and widely cited public dataset for smartphone-sensing depression research and is therefore a natural setting for a deliberately conservative re-analysis. The contribution of the present paper is not a new depression classifier; it is a baseline-controlled falsification attempt — does the missingness-as-signal hypothesis survive these four standards on a cohort that is already published and has been used as the basis of larger claims?

We address two complementary questions. First, between-person: does missingness predict post-term PHQ-9 or PHQ-9 change beyond pre-term PHQ-9? Second, within-person: does daily mood predict same-day or next-day continuous-sensor coverage? The two questions probe different timescales of the same hypothesised mechanism. We acknowledge upfront that the analyses were not pre-registered and are exploratory.

## Results

### Sample, missingness profile and effective dimensionality

The canonical StudentLife sample is the 48 undergraduates reported by Wang et al. 2014^6^. The public data archive, however, contains 59 distinct sensor-instrumented UIDs (numbered 0–60 with two gaps); the additional 11 are sparse-data records and PHQ-9 non-completers (characterised in detail in Methods, Data source) that contribute nothing to any between-person model in this paper, because all between-person analyses are restricted by an outcome-defined rule to the 46 UIDs with a pre-term PHQ-9 score (mean 5.5, s.d. 4.6, range 0–23; 17% scored ≥10) and the 38 with paired pre/post PHQ-9 (Table 1; CONSORT-style sample flow in Fig. 1). Mean post-term PHQ-9 was 6.3 (s.d. 5.8) and mean within-person change was +0.5 (s.d. 3.6); 7 of 38 (18%) exceeded the moderate-depression cutoff at post-term. The cohort thus experienced a mean PHQ-9 change smaller than the test’s typical short-interval reliability ^7^, implying that any predictor for change would have to discriminate within a small dynamic range.

**Table 1.** Sample and data summary. The canonical StudentLife sample is the 48 enrolled undergraduates reported by Wang et al. 2014; the public archive contains 59 distinct UIDs, an 11-UID excess characterised in Methods (Data source). All between-person analyses are restricted to the PHQ-9-defined subsamples (46 with pre-term PHQ-9, 38 with paired pre/post PHQ-9) and are therefore unaffected by the interpretation of the 11 additional UIDs. Continuous-sensor coverage is the proportion of participant-days with ≥ 1 record from any of the nine continuous streams.

| Characteristic | Value |
| --- | --- |
| Originally enrolled (Wang et al. 2014) | 48 |
| Distinct UIDs in public archive | 59 |
| Pre-term PHQ-9 administered | 46 |
| Post-term PHQ-9 administered | 38 |
| Both pre- and post-term (analytic sample) | 38 |
| Pre-term PHQ-9, mean (s.d.) | 5.52 (4.61) |
| Range | 0–23 |
| Pre PHQ-9 $\geq 10$, n (%) | 8 (17%) |
| Post-term PHQ-9, mean (s.d.) | 6.26 (5.84) |
| Range | 0–25 |
| Post PHQ-9 $\geq 10$, n (%) | 7 (18%) |
| PHQ-9 change (post – pre), mean (s.d.) | +0.45 (3.55) |
| Range | –8 to +10 |
| Observation window | 27 March – 31 May 2013 |
| Days observed | 66 |
| Continuous sensor streams | 9 |
| Phone-activity log streams | 5 |
| EMA prompt streams | 27 |
| Mean continuous-sensor coverage (any-stream day) | 96% |
| Mean per-stream day-level missingness, continuous | 22% |
| Mean per-stream day-level missingness, phone logs | 34% |
| Mean per-stream day-level missingness, EMA prompts | 90% |

**Table 2.** Joint and nested *F*-tests of missingness as an incremental predictor over pre-term PHQ-9. Permutation *P* from 2,000 within-row shuffles. The omnibus joint *F*-test of all nine continuous-stream day-level missingness rates against PHQ-9 (adjusted for pre PHQ-9) gave *F* (9, 27) = 0.43, *P* = 0.91.

| Outcome | Baseline $R^2$ | Full-model $R^2$ | Nested F (5 a-<br>priori features) | Parametric P | Permutation P (B<br>= 2,000) | Joint F (all 9<br>streams) | Joint P |
| --- | --- | --- | --- | --- | --- | --- | --- |
| Post-term PHQ-9 | 0.63 | 0.65 | $F(5,31) = 0.43$ | 0.821 | 0.818 | $F(9,27) = 0.43$ | 0.910 |
| $\Delta$ PHQ-9 | 0.00 | 0.07 | $F(5,31) = 0.43$ | 0.821 | 0.814 | $F(9,27) = 0.43$ | 0.910 |

**Fig. 1.**
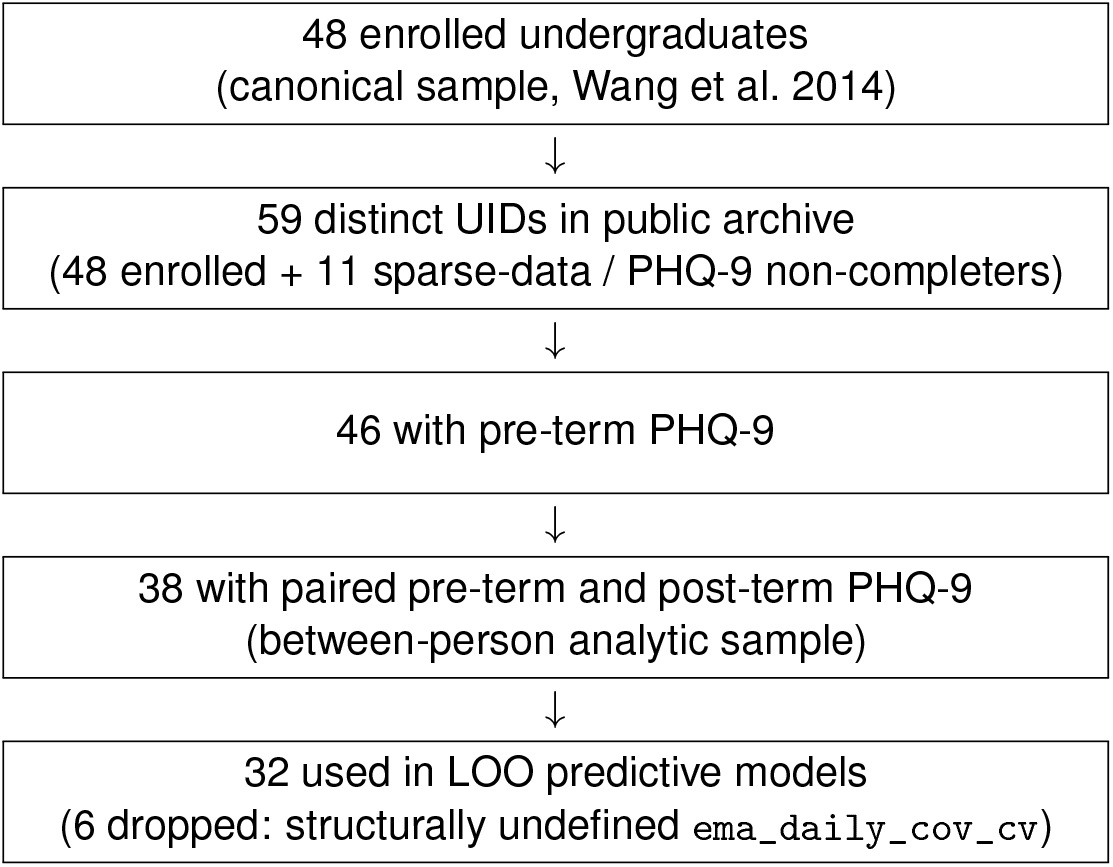
Sample-flow diagram. The canonical StudentLife sample is the 48 undergraduates reported by Wang et al. 2014. The public data archive contains 59 distinct sensor-instrumented UIDs (numbered 0–60 with two gaps); the 11-UID excess is heterogeneous (most produce only sparse single-stream data, e.g. Wi-Fi-only records with no PHQ-9 surveys, but at least one — UID 59 — has the highest PAM-EMA response count in the archive yet no pre-PHQ-9). The discrepancy is also reflected in the third-party `studentlife` CRAN package ^12^, which describes the dataset as 48 students while loading the full UID range. **All between-person analyses in this paper are restricted by an outcome-defined rule to the 46 UIDs with pre-term PHQ-9 and the 38 with paired pre/post PHQ-9; the 11 additional UIDs are by construction excluded from every between-person model and contribute only to the technical** 59 ×66× 41 **presence tensor before it is collapsed to PHQ-9 subsamples** (see Methods, Data source). The drop from 38 to 32 in the predictive sample is structural: six participants had a defined pre/post-term PHQ-9 but had a mean daily EMA-stream coverage of zero, which makes the EMA-derived feature `ema_daily_cov_cv` (coefficient of variation of daily EMA-stream coverage) undefined. The within-person day-level analysis used a separate sample of 2,186 person-days from 49 participants (no PHQ-9 filter applied).

Continuous-sensor coverage was high on most participant-days but uneven across streams (Fig. 2, Fig. 3). On 96% of participant-days at least one of the nine continuous streams produced a record. Mean per-stream day-level missingness was 22% across the continuous family and 34% across phone-activity logs; EMA-prompt streams had a mean of 90% missingness, but this combines true non-response with prompt-schedule sparsity. Cohort-mean continuous-sensor coverage drifted downward by ∼25 percentage points from late March to late May (Fig. 2b).

**Fig. 2.**
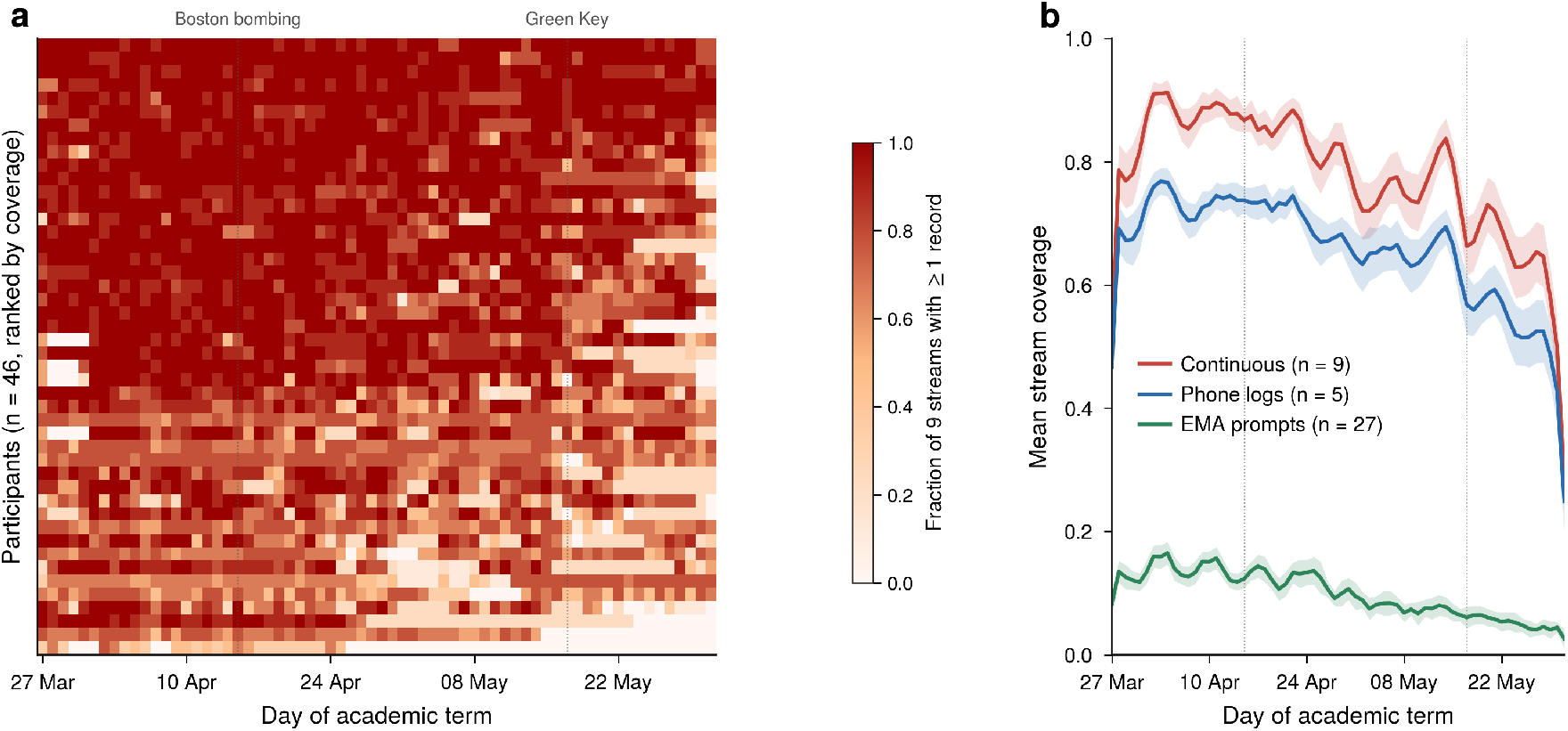
Continuous-sensor coverage across the StudentLife observation window. **a**, Per-participant per-day coverage matrix for the 46-person PHQ-9 cohort, sorted by mean coverage. Each row is one participant, each column one of 66 days; cell colour encodes the fraction of nine continuous-sensor streams producing ≥1 record on that day. **b**, Cohort-mean stream-coverage trajectories for the three stream families: continuous sensors (red, *n*_streams_ = 9), phone-activity logs (blue, *n*_streams_ = 5), EMA prompts (green, *n*_streams_ = 27). Lines show smoothed cohort means; shaded bands show ±1 s.e.m. (*n* = 46 participants). Vertical dotted lines mark the Boston Marathon bombing (15 April 2013) and the Dartmouth Green Key festival (17 May 2013). *Takeaway:* continuous-sensor coverage was high on most days but drifted downward by ∼ 25 percentage points over the term.

**Fig. 3.**
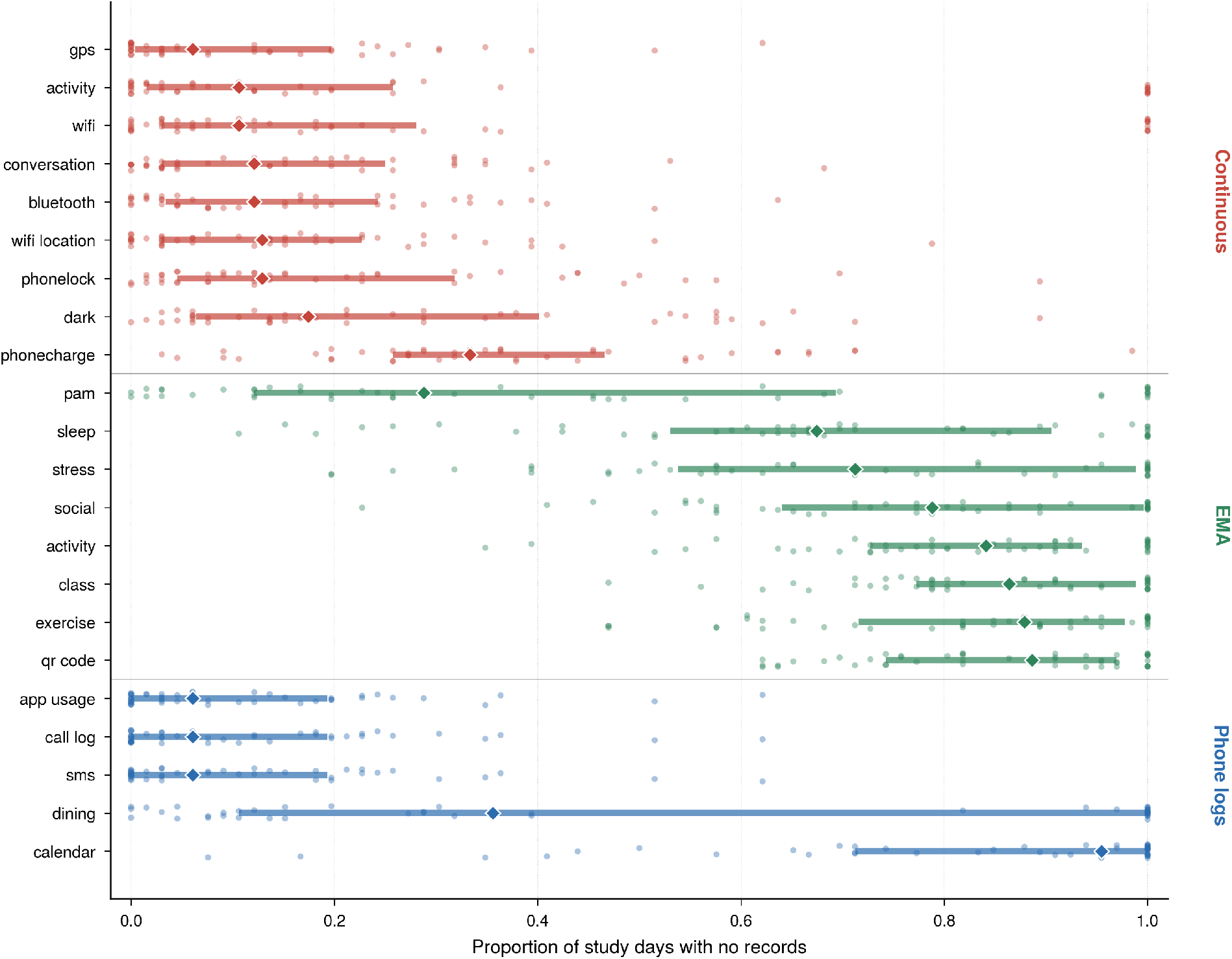
Per-stream day-level missingness across the 46-participant cohort. Each point is one participant; each row is one stream from one of three families: continuous sensors (red), EMA prompts (green), phone-activity logs (blue). Box-and-whisker style summaries indicate the cohort distribution of per-participant missingness (proportion of 66 study days with no records for that stream). EMA values combine prompt-schedule sparsity (most prompts triggered on a minority of days by design) with non-response and are not directly comparable to passive sensors that ran continuously. *Takeaway:* continuous sensors and phone-activity logs had per-participant median missingness ≲ 0.2; EMA streams had per-participant median missingness ≥ 0.7.

Although we computed 89 missingness features, principal-components analysis of the standardized feature matrix indicated substantial redundancy (Supplementary Table S1): two principal components captured 50% of the feature-set variance, seven captured 80%, and the Cheverud–Nyholt ^8^ effective number of independent dimensions was 5.5 (Kaiser eigenvalue ≥1: 12 components). Reported FDR-corrected *q* values across 89 features therefore overestimate the multiplicity correction by approximately a factor of 16 in independent-dimension terms; our headline qualitative findings (no feature significant after correction) are unchanged under either calibration.

### Baseline-adjusted associations

The strongest negative correlate of post-term PHQ-9 was the proportion of late-term days without any EMA response (Pearson *r* = − 0.37, *P* = 0.021, 95% bootstrap CI [−0.58, −0.10]; Fig. 4a,b). Several other EMA-engagement features fell in the same direction with similar magnitudes. After Benjamini–Hochberg FDR correction across all 89 features, no association survived (smallest *q* ≈0.50). Continuous-sensor missingness features did not correlate meaningfully with either outcome.

**Fig. 4.**
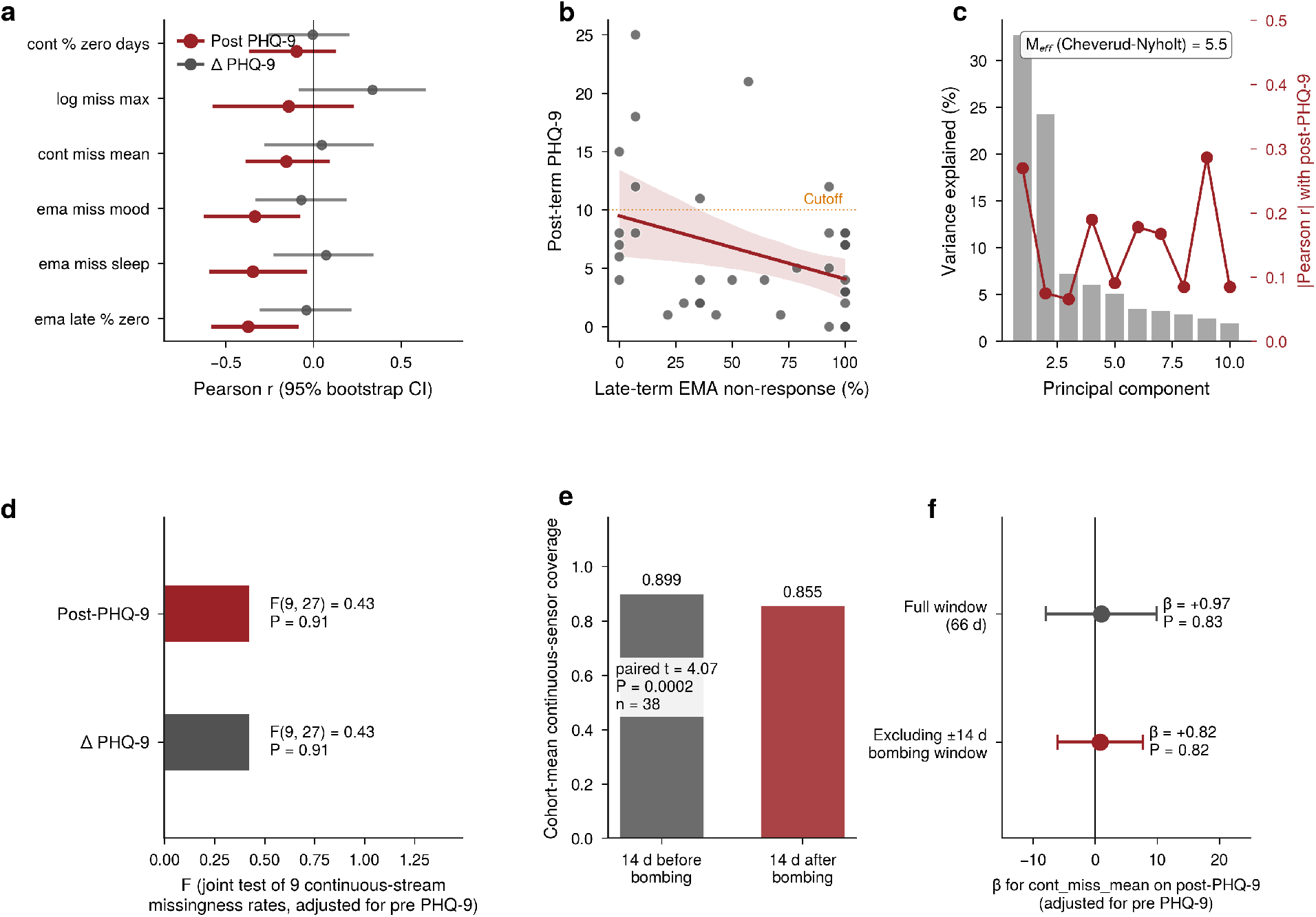
Between-person associations between missingness features and depression outcomes (*n* = 38). **a**, Bootstrap 95% CIs (*B* = 2,000) for the Pearson correlation between six representative missingness features and post-term PHQ-9 (red) or PHQ-9 change (grey). No association survived Benjamini–Hochberg multiplicity correction across the 89 features tested. **b**, Late-term EMA non-response versus post-term PHQ-9: the strongest nominally significant univariate relationship in the screen (*r* = −0.37, *P* = 0.021, *q* (BH) = 0.50). **c**, Principal-components analysis of the 89-feature missingness matrix (*M*_eff_ = 5.5 effective independent dimensions). **d**, Omnibus joint *F*-test of the nine continuous-stream day-level missingness rates against PHQ-9 outcomes adjusted for pre-PHQ-9 (*F* (9, 27) = 0.43, *P* = 0.91 for both outcomes). **e**, Cohort-mean continuous-sensor coverage in the 14-day windows before and after 15 April 2013 (paired *t* = 4.07, *P <* 0.001). **f**, OLS coefficient of mean across-stream continuous missingness on post-PHQ-9 (adjusted for pre-PHQ-9), with and without the ±14-day exclusion window — stable in direction, magnitude and significance. *Takeaway:* no individual missingness feature survived Benjamini–Hochberg multiplicity correction across the 89 features (panel a), and the omnibus joint *F*-test of the nine continuous-stream missingness rates against PHQ-9 was non-significant after baseline adjustment for pre-PHQ-9 (panels d, f). The between-person picture is robust to exclusion of the Boston Marathon bombing window. *Caution: exploratory, not pre-registered*.

As an omnibus test of whether continuous-sensor missingness rates were associated with depression outcomes after adjustment for baseline PHQ-9, we jointly tested the nine continuous-stream day-level missingness rates against post-term PHQ-9 in a nested *F*-framework. The joint test was non-significant (*F* (9, 27) = 0.43, *P* = 0.91; Fig. 4d). The same joint test for PHQ-9 change was identically non-significant. This provides no evidence in this cohort that the continuous-sensor missingness pattern depends on depression severity. A complementary OLS model regressing post-term PHQ-9 on five a priori continuous-sensor missingness features adjusted for pre-term PHQ-9 explained *R*^2^ = 0.66, driven essentially entirely by pre-term PHQ-9 (*β* = 0.91, 95% CI [0.63, 1.20], *P <* 0.001; Supplementary Table S2). The corresponding nested *F*-test against the pre-PHQ-9-only baseline was non-significant (*F* (5, 31) = 0.43, parametric *P* = 0.82, permutation *P* = 0.82 over 2,000 permutations; Δ*R*^2^ = 0.024).

Principal-components-based exploration was equally null. For post-term PHQ-9 the top |*r*| was 0.27 for PC1 (*P* = 0.10); for PHQ-9 change the smallest uncorrected *P* was 0.013 for PC6, but this did not survive Benjamini–Hochberg correction across the twenty PC–outcome tests (*q* ≈0.13). No PC–outcome correlation reached *q <* 0.05 in either outcome (Fig. 4c, Supplementary Table S3).

### Predictive performance under cross-validation

Leave-one-out cross-validation (LOO-CV) reproduced the regression picture under a stricter test. Pre-term PHQ-9 alone yielded LOO *R*^2^ = 0.59 for post-term PHQ-9 (95% cluster-bootstrap CI [0.22, 0.81]; Pearson *r* = 0.77, *P <* 0.001; Table 3). The bootstrap CI is wide, reflecting the small sample, but its lower bound at +0.22 still substantially exceeds the cohort-mean baseline (*R*^2^ = −0.07). All three regularized linear models trained on missingness features alone, with *α* and *ℓ*_1_-ratio chosen by 3-fold inner CV, gave LOO *R*^2^ between −0.08 and 0.23 — statistically indistinguishable from the cohort-mean baseline. A conservatively parameterized random forest (300 trees, min_samples_leaf = 5, 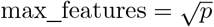, max_depth = 5) trained on missingness alone gave *R*^2^ = −0.10. When missingness was combined with pre PHQ-9 the random forest reached *R*^2^ = +0.02 — below the lower bound of the baseline CI ([0.22, 0.81]; Fig. 6a) and therefore not evidence of incremental signal. For PHQ-9 change (Fig. 6b), no model achieved positive LOO *R*^2^.

**Table 3.** Predictive performance of nested-CV-tuned models for post-term PHQ-9 and PHQ-9 change under leave-one-out cross-validation (*n* = 32). Pre PHQ-9 only LOO *R*^2^ = +0.59, with cluster-bootstrap 95% CI [0.22, 0.81].

| Model | Post $R^2$ | Post RMSE | $\Delta R^2$ | $\Delta$ RMSE |
| --- | --- | --- | --- | --- |
| Mean baseline | –0.07 | 6.30 | –0.07 | 3.78 |
| Pre PHQ-9 only | +0.59 | 3.93 | –0.15 | 3.93 |
| Lasso (miss only, tuned) | –0.21 | 6.70 | –0.07 | 3.78 |
| Ridge (miss only, tuned) | –0.08 | 6.34 | –0.11 | 3.85 |
| ElasticNet (miss only, tuned) | –0.23 | 6.76 | –0.07 | 3.78 |
| RF (miss only, conservative) | –0.10 | 6.40 | –0.19 | 4.00 |
| Ridge (miss + pre, tuned) | –0.07 | 6.32 | –0.11 | 3.86 |
| RF (miss + pre, conservative) | +0.02 | 6.03 | –0.20 | 4.01 |

For binary classification of post-term PHQ-9 ≥10 (Fig. 6c, Table 4), pre-term PHQ-9 alone gave a LOO area under the ROC curve (AUC) of 0.87 with bootstrap CI [0.69, 1.00]. The L2 logistic model on missingness alone gave AUC = 0.03 [0.00, 0.10] and the L1 logistic gave 0.25 [0.03, 0.47]; both bootstrap CIs lay entirely below 0.5. The conservatively parameterized random forest on missingness alone gave AUC = 0.41 [0.18, 0.67], with the bootstrap CI including 0.5. Combining missingness with pre PHQ-9 in a logistic model degraded the baseline AUC from 0.87 to 0.16 [0.00, 0.46]. Because the binary analysis contained only seven positive cases, AUC estimates and their bootstrap intervals should be interpreted as instability diagnostics for the missingness-as-signal hypothesis rather than as reliable classifier-performance estimates. Calibration of the LOO classifiers (Supplementary Fig. S4) confirmed that the pre-PHQ-9 baseline produced well-calibrated predicted probabilities (predicted 1%, 5%, 12%, 68% matched observed event rates of 0%, 14%, 11%, 63% across quartile bins), whereas the missingness-only L2 logistic was anti-calibrated (highest-predicted-probability quartile contained zero observed events; lowest-predicted-probability quartile contained 38% observed events). The inverted direction of the linear missingness-only AUCs is consistent between the OLS coefficient signs and the binary classification, but does not support the missingness-as-withdrawal interpretation in StudentLife and should be replicated before further inference.

**Table 4.** Classification of post-term PHQ-9 ≥10 (*n* = 32; 7 positive cases) under leave-one-out cross-validation, with bootstrap 95% AUC CIs (*B* = 2,000). Because the binary analysis contained only seven positive cases, AUC estimates and their bootstrap intervals should be interpreted as instability diagnostics rather than as reliable classifier-performance estimates.

| Model | n | AUC (95% bootstrap CI) | Direction at chance? |
| --- | --- | --- | --- |
| Pre PHQ-9 only | 32 | 0.874 [0.689, 1.000] | Excluded — significantly above chance |
| L2 logistic (miss, tuned) | 32 | 0.029 [0.000, 0.096] | Excluded — significantly below chance |
| L1 logistic (miss, tuned) | 32 | 0.249 [0.027, 0.474] | Excluded — significantly below chance |
| RF (miss, conservative) | 32 | 0.411 [0.178, 0.667] | CI includes chance |
| L2 logistic (miss + pre, tuned) | 32 | 0.160 [0.000, 0.455] | Excluded — significantly below chance |

To rule out the possibility that the absent missingness signal reflects a failure of the LOO pipeline rather than a property of the data, we ran a positive-control experiment in which fake biomarkers calibrated to deliver incremental *R*^2^ of 0.10, 0.20 and 0.30 above the pre-PHQ-9 baseline were injected as single columns. The pipeline recovered the injected signals to within ∼10% of target (+0.086, +0.191, +0.297 across 50 random seeds; Supplementary Fig. S2). The pipeline is therefore well-calibrated to detect incremental signals at the magnitudes that would be clinically interesting, and the observed null is a feature of the data, not the analysis. A complementary feature-engineering ablation (Supplementary Fig. S3, Supplementary Table S7) re-ran the LOO ridge under six alternative parameterizations of the missingness feature-set — continuous-stream-only, phone-log-only, EMA-only, aggregate cohort-level-summary block (eight summary types × three stream families = 24 features), PCA top-five rotation, and the manuscript’s primary all-89-feature set — and showed that no parameterization yielded miss-only LOO *R*^2^ *>* 0 or miss-plus-pre LOO *R*^2^ exceeding the pre-PHQ-9-only baseline by more than 0.01. The closest competitor was the aggregate-summary block (*R*^2^ = +0.592 vs +0.585 for pre alone, Δ = +0.006), which is the cleanest possible demonstration of the absence of incremental signal.

### Sensitivity to the Boston Marathon bombing window

We assessed whether the Boston Marathon bombing (15 April 2013) confounded the analysis. Cohort-mean continuous-sensor coverage dropped significantly from the 14-day pre-bombing window (mean 0.899) to the ±14-day post-bombing window (mean 0.855; paired *t* = 4.07, *P <* 0.001; Fig. 4e). However, the OLS coefficient of mean across-stream continuous missingness on post-term PHQ-9 (adjusted for pre-PHQ-9) was stable in direction, sign and significance when the ±14-day window around the bombing was excluded (*β*: +0.97 →+0.82, both *P >* 0.8; Fig. 4f, Supplementary Table S4). The bombing produced a real cohort-wide coverage drop but did not change the missingness–PHQ-9 association we observed.

### Within-person mood and next-day coverage

Because the Photographic Affect Meter (PAM ^9^) is a 4×4 valence × arousal grid rather than a unidimensional mood scale, we decomposed each PAM response into its valence component (column index, 1–4) and arousal component (row index, 1–4) before within-person *z*-scoring. We then merged daily mood with daily continuous-sensor coverage (mean of nine streams) to obtain 2,186 person-day pairs across 49 participants. The composite (raw picture-index) within-person correlation with same-day coverage was small and not statistically distinguishable from zero (cluster-bootstrap *r* = +0.032, 95% CI [−0.025, +0.089]; Fig. 5a, b, Table 5).

**Table 5.** Within-person daily mood vs continuous-sensor coverage: composite, valence and arousal. Pearson *r* computed within-person after demeaning each variable within participant; 95% CI from cluster-bootstrap over participants (*B* = 1,000). 2,186 person-days across 49 participants. Inference uses the cluster-bootstrap CI (reflecting clustering by participant); we do not report parametric *P* -values because tests that treat the 2,186 person-days as independent are anti-conservative under within-participant clustering (ICC = 0.42).

| Mood dimension | Lag | $r$ | 95% cluster-bootstrap CI | CI excludes 0? |
| --- | --- | --- | --- | --- |
| Composite (z-scored picture_idx) | same-day | +0.032 | [−0.025, +0.089] | No |
| Composite (z-scored picture_idx) | next-day | −0.010 | [−0.063, +0.054] | No |
| Valence | same-day | +0.061 | [−0.007, +0.136] | No |
| Valence | next-day | +0.082 | [+0.011, +0.162] | Yes |
| Arousal | same-day | +0.019 | [−0.023, +0.064] | No |
| Arousal | next-day | −0.031 | [−0.078, +0.018] | No |

**Fig. 5.**
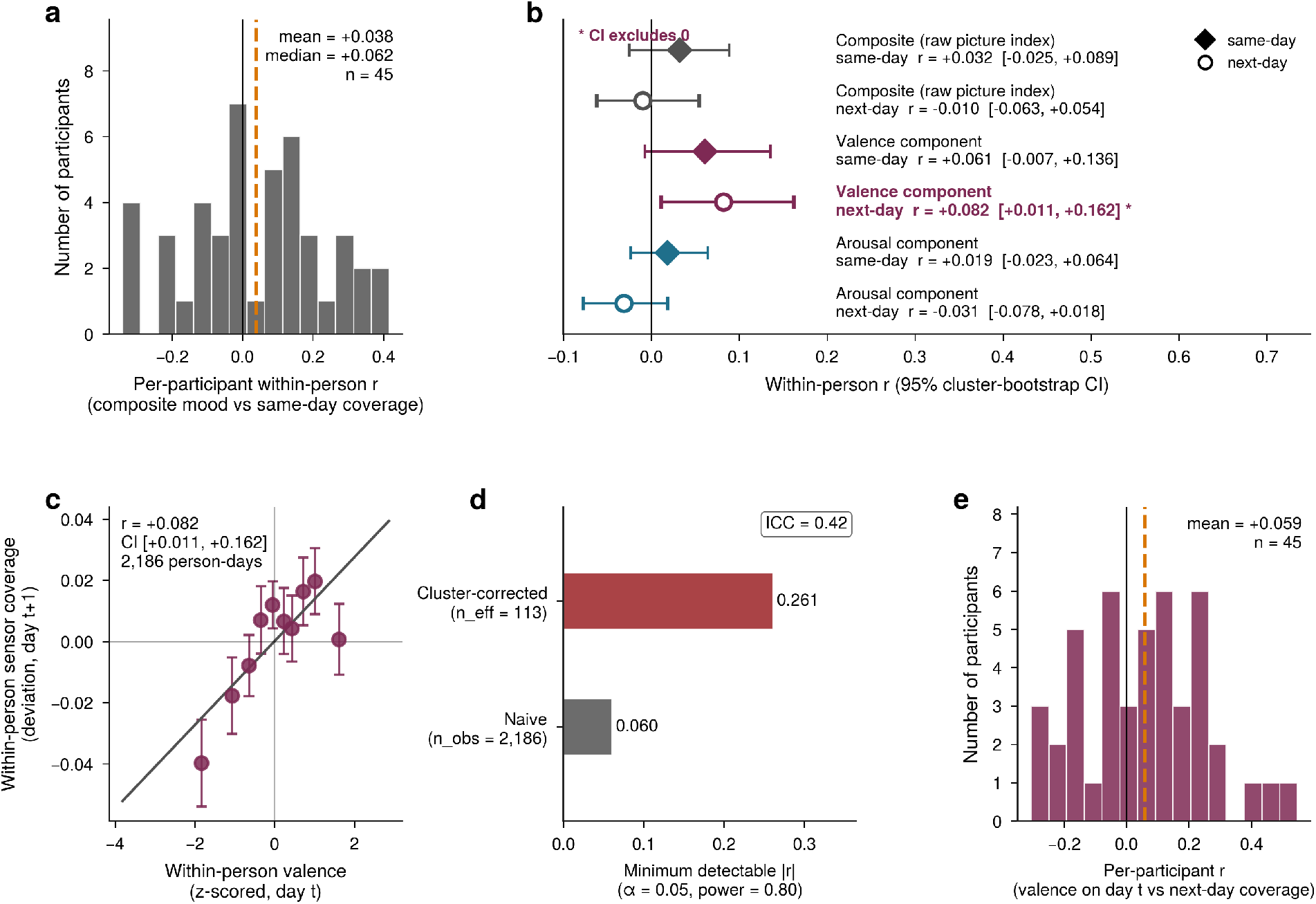
Within-person daily mood and sensor-coverage missingness (2,186 person-days from 49 participants for the pooled analyses; per-participant correlations in panels **a** and **e** are shown for the 45 participants with ≥ 5 valid PAM–coverage day pairs after demeaning, since fewer pairs make a within-person Pearson *r* undefined or singular). **a**, Distribution of per-participant Pearson correlations between within-person daily PAM composite (raw picture index) and same-day continuous-sensor coverage. **b**, Pooled within-person *r* (cluster-bootstrap 95% CIs) for composite, valence and arousal mood components against same-day and next-day coverage. The valence next-day correlation (*r* = +0.082, 95% CI [+0.011, +0.162]) is the only correlation whose CI excludes zero, in the direction *opposite* to the withdrawal hypothesis. **c**, Pooled within-person association between valence on day *t* and continuous-sensor coverage on day *t* + 1 (binned). **d**, Power analysis: ICC = 0.42, *n*_eff_ = 113, minimum detectable |*r*| = 0.26 clustered. **e**, Distribution of per-participant valence-next-day correlations. *Takeaway:* the within-person valence effect is small, prospective, opposite in direction to the withdrawal hypothesis, and only marginally powered. *Caution: exploratory, not pre-registered*.

**Fig. 6.**
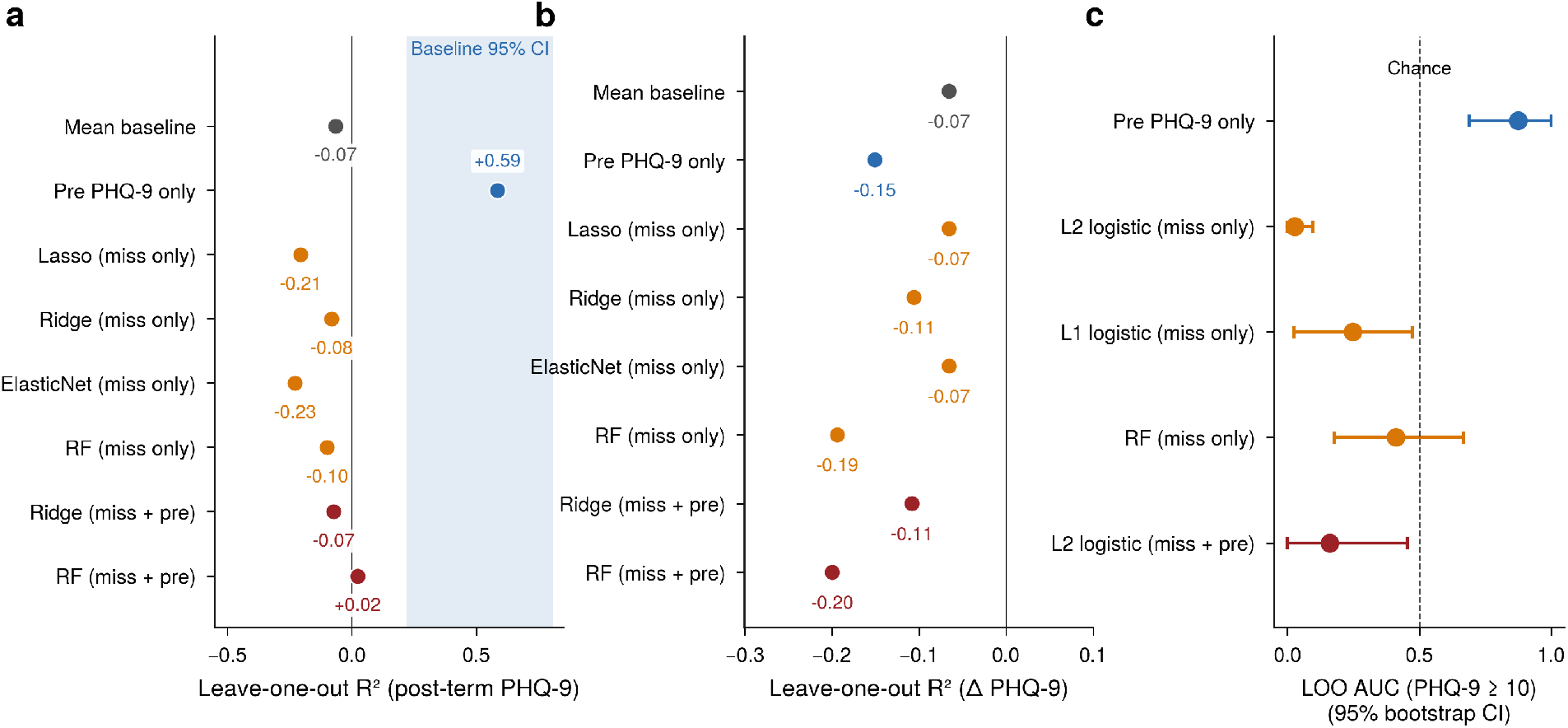
Predictive utility of missingness features under leave-one-out cross-validation (*n* = 32), with nested-CV-tuned hyperparameters. **a**, LOO *R*^2^ for each model predicting post-term PHQ-9. The shaded blue band is the cluster-bootstrap 95% CI of the pre-PHQ-9-only baseline ([0.22, 0.81]). All tuned linear models on missingness alone fall in the cohort-mean baseline region; the random-forest miss+pre point estimate (+0.02) sits below the lower bound of the baseline CI. **b**, Same comparison for predicting PHQ-9 change. **c**, LOO area under the ROC curve for classifying post-term PHQ-9 ≥10 (7 positive cases), with 95% bootstrap CIs (*B* = 2,000). The conservatively parameterized random-forest CI includes 0.5; linear missingness-only CIs lie entirely below 0.5. *Takeaway:* no missingness model improves on the baseline self-report. *Caution: AUC values should be interpreted as instability diagnostics rather than as reliable classifier-performance estimates because the positive class contains only 7 cases*.

Decomposition revealed a difference between dimensions. The valence-component cluster-bootstrap correlation with next-day coverage was *r* = +0.082, 95% CI [+0.011, +0.162], with the CI excluding zero (Fig. 5b, c). The same-day valence correlation was *r* = +0.061, 95% CI [−0.007, +0.136], borderline. Both arousal correlations (same-day *r* = +0.019; next-day *r* = −0.031) had CIs that included zero. The direction of the valence effect is opposite to the withdrawal-equals-missing-data hypothesis: in these data, days on which a participant rated their mood more positively were followed by more, not less, continuous-sensor data. The effect is small (*r* ≈0.08, equivalent to ∼0.7% of within-person coverage variance) and we caution against strong interpretation given that the analyses were exploratory and not pre-registered. With 2,186 observations clustered within 49 participants and an ICC of 0.42, the design effect implies an effective sample size of *n*_eff_ = 113 and a minimum detectable |*r*| of 0.26 at *α* = 0.05 and power = 0.80. The within-person analysis is well-powered to rule out medium effects and only marginally powered against small effects.

## Discussion

In the StudentLife cohort, smartphone-data missingness did not add incremental predictive value for post-term PHQ-9 beyond a single baseline self-report. Univariate associations did not survive multiplicity correction; nested-CV-tuned regularized linear models trained on missingness alone matched the cohort-mean baseline; an omnibus joint *F*-test of all nine continuous-stream missingness rates against PHQ-9, adjusted for pre-PHQ-9, was non-significant. A separately motivated within-person day-level analysis revealed a small valence-specific prospective effect (*r*≈ +0.08) opposite in direction to the withdrawal hypothesis, with arousal null.

A single pre-term PHQ-9 is a remarkably hard benchmark to beat for end-of-term PHQ-9 (LOO AUC 0.87 for the clinical cutoff) because depression in this cohort is largely stable on a ten-week timescale: the test-retest correlation across the term is high and the within-person change is small. This means that any candidate digital biomarker added to a model also containing the baseline self-report has a small remaining variance to explain, and the comparison to a baseline-only model is the relevant test of incremental signal. Many published passive-sensing depression analyses report performance only against a cohort-mean baseline, which can give an inflated picture of contribution.

This analysis should not be read as evidence that missingness is never informative. It shows that, in a high-functioning undergraduate cohort over a ten-week term, missingness did not add incremental predictive value beyond a single baseline PHQ-9. In clinical samples, in longer observation horizons, or in populations with greater dynamic range in symptom severity and engagement, dropout has more room to track psychiatric state ^10,11^. The conservative null reported here is a constraint on what can be claimed from this dataset, not a universal claim about smartphone-data missingness in mental health.

The between-person and within-person analyses answer different questions. The originating withdrawal hypothesis is a slow, between-person process: depressed individuals withdraw from device wear over weeks. The within-person analysis instead tests state-dependent engagement — whether a person’s daily mood predicts their daily-to-following-day sensor coverage. A null on the latter does not exonerate the former, and the small valence effect we observed is noteworthy because it inverts the prevailing intuition: better mood today predicts more, not less, sensor data tomorrow. One mechanistic reading is that more positive mood reflects engagement with the social and academic rhythms that incidentally produce more sensor records (more conversation, more movement, more app usage); a darker reading is that EMA-engagement and sensor-coverage features partially proxy for conscientiousness, social embedding or institutional anchoring, all of which are themselves protective factors and which the digital-phenotyping literature has documented as correlates of retention in remote mental-health studies ^3,4^. Disentangling these mechanisms requires designs that did not exist in the StudentLife protocol.

Methodologically, our results illustrate three recurring failure modes in small-sample digital phenotyping that careful baseline comparison brings into focus. (i) When 89 features are matched against an outcome with effective *n* = 38, even nested-CV-tuned regularized linear models can only match the cohort-mean baseline; the question is not whether they outperform that baseline but whether they outperform the baseline self-report.(ii)Headline feature counts can substantially overstate effective dimensionality: the 89 features in our analysis carry approximately 5.5 effective independent dimensions. FDR correction over the nominal feature count is therefore overconservative, and reports of high-dimensional missingness signatures should distinguish nominal from effective dimensionality. (iii) Univariate screens followed by selective reporting can produce nominally significant associations that do not survive an omnibus test on the same data. We recommend a small set of reporting standards (Box 1) before missingness is interpreted as signal.

This study has several important limitations. The analytic between-person sample is small (*n* = 38), and the cluster-bootstrap CI on the baseline LOO *R*^2^ is correspondingly wide ([0.22, 0.81]) — the cohort cannot rule out missingness effects of moderate size. A Monte Carlo power simulation under the empirical predictor distribution (Supplementary Section S7, Supplementary Fig. S1, Supplementary Table S6) shows that the joint *F*-test reached 80% power at an incremental *R*^2^ of 0.17 above the pre-PHQ-9 baseline; the cohort therefore could detect large incremental missingness signals but could not rule out small-to-moderate ones (power 22% at Δ*R*^2^ = 0.05; 46% at Δ*R*^2^ = 0.10). StudentLife is an Android-based, undergraduate-only dataset from spring 2013 with limited clinical severity and limited PHQ-9 change; results may not generalise to clinical samples, to current-generation operating systems and apps, or to older populations. The analyses were exploratory and were not pre-registered; effect sizes and directions reported here do not constitute confirmatory evidence and should be replicated with pre-registered hypotheses in larger and more clinically heterogeneous datasets.

EMA-prompt missingness conflates true non-response with prompt-schedule sparsity. Binary classification was constrained by only seven positive cases; the AUC bootstrap intervals reported here should be interpreted as instability diagnostics, not as reliable estimates of classifier performance. The predictive sample of *n* = 32 slightly over-represents EMA-engaged participants relative to the *n* = 38 between-person sample because six participants had structurally undefined values for one EMA-derived feature; these six had lower mean PHQ-9 (pre 3.2 vs 6.3; post 4.8 vs 6.5), and a multiple-imputation sensitivity analysis on *n* = 38 (MICE-style `IterativeImputer`with Bayesian-ridge regression, *m* = 20; Supplementary Table S7) leaves the headline LOO *R*^2^ for pre-PHQ-9 essentially unchanged (0.58–0.59) and confirms no incremental signal from missingness.

In a baseline-controlled, cross-validated, multiplicity-aware re-analysis of the StudentLife cohort, smartphone-data missingness did not add incremental predictive value for depression beyond a single baseline PHQ-9. The result is exploratory and StudentLife-specific. Its practical implication is narrower than a universal claim: missingness should not be interpreted as psychological signal in the absence of baseline-controlled, cross-validated, multiplicity-aware evidence.

### Box 1. Recommended reporting standards before interpreting smartphone-data missingness as psychological signal

- Report missingness models with and without baseline self-report on the same outcome under cross-validation.
- Use nested cross-validation (or held-out tuning sets) for any model with tuned hyperparameters.
- Report omnibus association tests (e.g., joint *F*, MANOVA), not only per-feature univariate screens.
- Apply multiplicity control (e.g., Benjamini–Hochberg FDR) across all tested features.
- Report effective dimensionality of the feature set (e.g., Cheverud–Nyholt *M*_eff_), not only the nominal feature count.
- Distinguish between-person and within-person missingness hypotheses, which probe different timescales of the proposed mechanism.
- Treat small-sample binary classification (e.g., positive class *<* 10) as instability diagnostics rather than performance estimates.
- Demonstrate pipeline calibration with a positive control (e.g., inject a synthetic biomarker calibrated to a known incremental *R*^2^ and confirm recovery) so that null findings can be distinguished from pipeline failure.
- Probe robustness to feature-engineering choices via an ablation across alternative parameterizations of the same underlying signal.
- For binary outcomes, report calibration curves (predicted vs observed event rates), not just AUC.

## Methods

### Pre-registration and analytic stance

The analyses reported here were not pre-registered. They are exploratory and were planned during a single re-analysis of the public dataset. Effect sizes and directions reported in this paper therefore do not constitute confirmatory evidence and should be replicated with pre-registered hypotheses in larger and more clinically heterogeneous samples.

### Data source

We used the publicly available StudentLife dataset ^6^. The original 2014 publication reports *n* = 48 enrolled undergraduates carrying instrumented Android smartphones for ∼10 weeks at Dartmouth College in Spring 2013. The public data archive contains 59 distinct sensor-instrumented UIDs (numbered 0–60 with two gaps), an 11-UID excess over the canonical Wang 2014 sample size. We resolved this discrepancy with a single, transparent rule: all between-person analyses are restricted to the 46 UIDs with a pre-term PHQ-9 score and the 38 UIDs with paired pre/post PHQ-9. The 11 additional UIDs are by construction excluded from every between-person model, correlation, *F*-test, AUC, calibration plot and incremental-*R*^2^ comparison reported in the paper, because none of those quantities is computable on a UID without PHQ-9 data. They appear only in the technical 59 × 66 × 41 presence tensor (Methods, Missingness feature engineering), which is then collapsed to the PHQ-9 subsamples before any analysis. For completeness, we inspected the per-UID data profiles: the additional UIDs are heterogeneous. Most produced only sparse data on a single stream (e.g. Wi-Fi scans only, with no other continuous-sensor records and no PHQ-9 surveys), consistent with test devices, partially-onboarded participants or devices that registered briefly before being removed from the study; a small number produced substantial sensor data — including UID 59, which has the highest PAM-EMA response count in the public archive — but never completed a pre-PHQ-9 survey, consistent with non-completion of self-report rather than absence from the cohort. The 11-UID excess is not described explicitly in the public dataset documentation; the independently developed CRAN package `studentlife`^12^ likewise treats the canonical sample as 48 students while loading the full UID range from the public archive, mirroring the same discrepancy. Our re-analysis is observational and used only the de-identified public release; no new data were collected, and no identifiable private information was generated, accessed or held by the author. This re-analysis qualifies as exempt human-subjects research under U.S. federal regulations governing secondary research use of publicly available, de-identified data (45 CFR §46.104(d)(4)). Original participant consent and de-identification were obtained and performed by the StudentLife investigators prior to public release of the dataset.

### Outcome and study window

The primary outcome was the Patient Health Questionnaire-9 (PHQ-9^7^). We scored items 1–9 on the standard 0–3 scale (range 0–27); item 10 was not used. Binary classification used a post-term cutoff of PHQ-9 ≥10. We restricted all missingness computations to a fixed 66-day window from 27 March 2013 to 31 May 2013.

### Sensor and EMA streams

We grouped 41 streams into three families. Continuous sensors (*n* = 9): activity inferences from the accelerometer, audio-derived conversation episodes, GPS fixes, Bluetooth scans, Wi-Fi scans, Wi-Fi-derived location bins, screen-dark events, phone-lock events and phone-charge events. Phone-activity logs (*n* = 5): foreground app-usage events, call log entries, SMS log entries, calendar entries and dining-card swipes. EMA prompts (*n* = 27) including the structured Photographic Affect Meter (PAM ^9^) and a battery of self-report mini-surveys. EMA-prompt missingness conflates true non-response with prompt-schedule sparsity (most prompts were triggered on a minority of days by design); we report EMA missingness on the same axis as continuous-sensor missingness in Fig. 3 but annotate this distinction explicitly.

### Missingness feature engineering and effective dimensionality

For each (participant, stream, day) cell we computed the number of records produced and a binary indicator of whether the stream produced at least one record on that day, yielding a 59 × 66 × 41 presence tensor. From this tensor we derived 89 participant-level features in three blocks. To estimate effective feature-set dimensionality we standardized the feature matrix, computed its eigenvalues, and reported the number of components needed to capture 50%, 80% and 95% of variance; the Kaiser eigenvalue ≥1count; and the Cheverud–Nyholt ^8^ effective number of independent tests 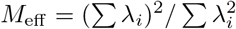.

### Statistical analysis

#### Univariate

Pearson and Spearman correlations between every missingness feature and each outcome, with Benjamini–Hochberg FDR control ^13^ across the 89 features. Bootstrap 95% confidence intervals (*B* = 2,000) for the top correlates by resampling participants with replacement.

#### Multivariable

OLS regression of each continuous outcome on five a priori continuous-sensor missingness features (`cont_miss_mean`, the cohort-mean across-stream missingness rate; `cont_pct_zero_days`, the proportion of days with zero continuous-stream coverage; `cont_longest_gap`, the longest consecutive run of days with zero continuous-stream coverage; `cont_cov_slope`, the OLS slope of daily continuous-stream coverage against day-of-term; and `cont_late_pct_zero`, the proportion of days in the last third of the term with zero continuous-stream coverage) adjusted for pre-term PHQ-9, with a nested *F*-test for incremental variance over a pre-PHQ-9-only baseline. Confirmed via 2,000-permutation test shuffling missingness predictors within rows. As an omnibus association test, joint *F*-test across all nine continuous-stream day-level missingness rates against each outcome adjusted for pre-PHQ-9.

#### Predictive

Leave-one-out cross-validation (LOO-CV) of: cohort-mean baseline, pre-term-PHQ-9-only baseline, lasso, ridge and elastic net (with *α* and *ℓ*_1_-ratio tuned on the training fold via 3-fold nested crossvalidation over a 7-point logarithmic grid for *α* and {0.2, 0.5, 0.8} for *l*_1_-ratio), and a random forest with conservative hyperparameters (300 trees, min_samples_leaf = 5, 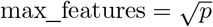, max_depth = 5). LOO *R*^2^, RMSE, MAE, Pearson *r* between predictions and observations, and bootstrap 95% CIs (*B* = 2,000) for the binary AUC. Cluster-bootstrap 95% CIs for the LOO *R*^2^ of the baseline ridge model (*B* = 200, resampling participants). The predictive sample contained *n* = 32 rather than *n* = 38 because one EMA-derived feature (*ema*_*daily* _*cov* _*cv*, the coefficient of variation of daily EMA-stream coverage) is structurally undefined for participants whose mean daily EMA-stream coverage is zero (six participants).

#### Boston-bombing sensitivity

Re-fit the OLS model after excluding participant-day observations within ±14 days of 15 April 2013. Paired *t*-test of cohort-mean continuous-sensor coverage in the 14-day windows before and after 15 April 2013.

#### Within-person day-level analysis

We loaded all PAM responses, decomposed each picture-grid index into its valence (column, 1–4) and arousal (row, 1–4) components per the original PAM specification ^9^, aggregated to the participant-day mean, and *z*-scored within participant. We merged with daily continuous-sensor coverage (the mean of nine binary stream-presence indicators) and computed within-person Pearson correlations after demeaning each variable within participant. Cluster-bootstrap 95% CIs were obtained by resampling participants with replacement (*B* = 1,000). The resulting analysis used 2,186 person-day observations across 49 participants.

#### Power calculation

We computed the within-person ICC of daily sensor coverage (ICC = 0.42), the design effect 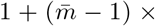 ICC for a mean cluster size 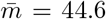, and the corresponding effective sample size *n*_eff_ = 113. The minimum detectable |*r*| at *α* = 0.05 (two-sided) and power = 0.80 was computed via Fisher’s *z*-transform.

#### Software

All analyses were conducted in Python 3 using NumPy, Pandas, SciPy and scikit-learn ^14^. The .Rds files were parsed in Python via a custom XDR-format serialization parser (described in the Supplementary Information).

## Data availability

All data analyzed in this study are from the publicly available StudentLife dataset (https://studentlife.cs.dartmouth.edu/dataset.html). No new data were collected. The pre-processed participant-day-stream presence tensor and the 89-feature participant-level table are included as supplementary files.

## Code availability

All analysis code is available as supplementary material and will be archived on Zenodo with a permanent DOI on acceptance. The code is written entirely in Python and reproduces all numerical results, tables and figures in approximately 15 minutes on a 2024 commodity laptop.

## Reproducibility

All numerical results are deterministic given the released code and the public dataset. Random-seed values are explicit in the code (seed = 0 for the primary bootstrap; seeds = 1, 2 for sensitivity bootstraps).

## Author contributions

C.O. is the sole author and is responsible for the conception, analysis and writing of this work.

## Acknowledgements

We thank the StudentLife team and the participants of the original 2013 study, whose data made this re-analysis possible. This re-analysis received no external funding.

## Supplementary Information

### S1. Effective dimensionality of the missingness feature matrix

Principal-components analysis of the standardized 89 × 38 missingness feature matrix is summarised in Supplementary Table S1. Components needed to capture 50%, 80% and 95% of variance: 2, 7 and 16 respectively. Kaiser (eigenvalue ≥ 1) component count: 12. Cheverud–Nyholt effective number of independent tests: 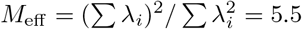

### S2. Multivariable OLS coefficients

Detailed coefficient tables for the OLS models of post-term PHQ-9 and PHQ-9 change on five a priori continuous-sensor missingness features adjusted for pre-term PHQ-9 are provided in Supplementary Table S2.

### S3. Principal-components correlations with PHQ-9

Pearson correlations between PC1–PC10 scores and PHQ-9 outcomes (post-term and change) are provided in Supplementary Table S3. The smallest uncorrected *P* across all twenty PC–outcome tests is *P* = 0.013 (PC6 vs Δ PHQ-9), which does not survive Benjamini–Hochberg correction across the twenty tests (*q* ≈0.13). No PC–outcome correlation reached *q <* 0.05 in either outcome.

### S4. Boston-bombing sensitivity analysis

Detailed sensitivity-analysis results, including pre/post window means and the OLS coefficient with and without the ±14-day exclusion window, are provided in Supplementary Table S4.

### S5. Custom RDS parser

The publicly released StudentLife data files are stored as R serialized .Rds files. The .Rds files were parsed in pure Python via a custom XDR-format serialization parser supporting the data types used in the StudentLife release (INTSXP, REALSXP, STRSXP, LGLSXP, RAWSXP, CHARSXP and VECSXP, with class attributes for data.frame, factor and POSIXct; and ALTREP-encoded compact integer/real sequences). The parser is included in the released analysis code.

### S6. Sensitivity analyses by baseline severity

We re-ran the top univariate associations stratified by pre-term PHQ-9 severity (cutoffs 5 and 10). No association strengthened materially within either stratum; the high-severity stratum at cutoff 10 contained only seven participants and was statistically uninformative.

### S7. Monte Carlo power simulation for the joint F-test

To quantify what missingness effects the joint *F*-test could plausibly have detected at *n* = 38, we ran a Monte Carlo power simulation under the actual cohort design. We took the observed pre-term PHQ-9 distribution and the observed 38 × 9 matrix of continuous-stream day-level missingness rates as fixed; estimated the null parameters of the post-PHQ-9-on-pre-PHQ-9 OLS model on the actual data 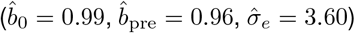; and at each target incremental *R*^2^ in {0, 0.025, 0.05, 0.075, 0.10, 0.125, 0.15, 0.175, 0.20, 0.25, 0.30, 0.40, 0.50}generated 1,000 synthetic post-PHQ-9 vectors with a planted incremental signal pointing in a random direction in the standardized 9-dimensional missingness-rate space, calibrated analytically to the target incremental *R*^2^. We then ran the same nested *F*-test the manuscript reports (full vs reduced; Methods →Statistical analysis→ Multivariable) and recorded the rejection rate at *α* = 0.05. Type-I error at Δ*R*^2^ = 0 was 0.045, very close to the nominal 0.05, confirming correct test calibration. The resulting power curve (Supplementary Fig. S1, Supplementary Table S6) crosses 80% power at incremental *R*^2^ = 0.17 above the pre-PHQ-9 baseline. The observed null in StudentLife (*F* (9, 27) = 0.43, *P* = 0.91) is therefore informative against missingness signals that would add 17 percentage points or more of incremental out-of-sample variance over a baseline self-report, but cannot rule out small-to-moderate incremental signals (e.g., Δ*R*^2^ = 0.05, where power is only 22%; or Δ*R*^2^ = 0.10, where power is 46%). Detecting effects of those magnitudes reliably at the same *α* would require samples several times larger than StudentLife.

### S8. Imputation sensitivity for the predictive sample

The LOO predictive analysis used *n* = 32 rather than *n* = 38 because the feature `ema_daily_cov_cv`(coefficient of variation of daily EMA-stream coverage) is undefined for six participants whose mean daily EMA-stream coverage is zero. To test whether the structural drop biased the headline result, we ran the headline ridge model (*Ridge (miss + pre)* from Table 3) on the full *n* = 38 under multiple imputation (MICE-style `IterativeImputer`with Bayesian-ridge per-feature regression and posterior sampling, *m* = 20 imputations averaged after pooling). The pre-PHQ-9-only baseline reaches LOO *R*^2^ = +0.579 on *n* = 38, essentially identical to the +0.59 reported for *n* = 32. The Ridge (miss + pre) model on *n* = 38 reaches LOO *R*^2^ = −0.012 under multiple imputation (sd = 0.062 across *m* = 20, range [−0.092, +0.111]), close to the published complete-case value of −0.07 and well below the pre-PHQ-9-only baseline (Supplementary Table S7). The headline finding (no incremental missingness signal beyond pre-PHQ-9) is robust to how the structural drop is handled.

### S9. Positive-control experiment for the LOO pipeline

To rule out the possibility that the absent missingness signal in Supplementary Fig. S1 reflects a failure of the LOO pipeline rather than a property of the data, we ran a positive-control experiment. We constructed a fake biomarker as a single column 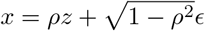, where *z* is the cross-validated residual of post-PHQ-9 after pre-PHQ-9, *ϵ* is independent standard Gaussian noise, and *ρ* is calibrated analytically so that adding *x* to a pre-PHQ-9-only ridge yields a target incremental LOO *R*^2^. We tested three target levels (0.10, 0.20, 0.30) over 50 random seeds each, and ran the same LOO ridge analysis the manuscript reports as primary. Observed mean incremental *R*^2^ was +0.086 (SD 0.050), +0.191 (SD 0.039), and +0.297 (SD 0.018) at the three target levels (Supplementary Fig. S2; Supplementary Table S8), recovering injected signals to within ∼10% of target. The pipeline is well-calibrated to detect incremental signals at the magnitudes that would be clinically interesting; the observed null in StudentLife is a feature of the data, not the analysis.

### S10. Feature-engineering ablation

We re-ran the headline LOO ridge analysis under six alternative parameterizations of the missingness feature-set: the manuscript’s primary all-89-feature set; continuous-stream-only (*p* = 25); phone-log-only (*p* = 21); EMA-only (*p* = 43); an aggregate cohort-level-summary block (*p* = 24, comprising eight summary types × three stream families: cohort-mean, max, min, s.d., percentage-of-zero-days, longest-gap, coverage-slope, late-percentage-zero, applied to the continuous, phone-log and EMA families separately); and a PCA top-five rotation of the standardized 89-feature matrix. Across all six parameterizations, miss-only LOO *R*^2^ remained negative and miss-plus-pre LOO *R*^2^ never exceeded the pre-PHQ-9-only baseline (LOO *R*^2^ = +0.585) by more than 0.01 (Supplementary Fig. S3, Supplementary Table S9). The closest competitor was the aggregate-summary block at miss-plus-pre LOO *R*^2^ = +0.592 (Δ = +0.006), which is the cleanest possible demonstration of the absence of incremental signal: a low-dimensional, interpretable missingness summary added almost exactly nothing to a single self-report when both were used together.

### S11. Calibration of LOO classifiers for the binary outcome

For binary classification of post-term PHQ-9 ≥10 (*n* = 32, 7 positive cases), we computed quartile-binned reliability curves for the LOO predicted probabilities of two models: a logistic regression on pre-PHQ-9 alone, and an L2 logistic on the full 89-feature missingness block (Supplementary Fig. S4, Supplementary Table S10). The pre-PHQ-9 baseline was reasonably well-calibrated: predicted probability quartile means of 0.01, 0.05, 0.12 and 0.68 corresponded to observed event rates of 0.00, 0.14, 0.11 and 0.63 respectively. The missingness-only L2 logistic was anti-calibrated: the highest-predicted-probability quartile (predicted mean 0.75) contained zero observed events, while the lowest-predicted-probability quartile (predicted mean 0.01) contained 38% observed events. This is a direct visualisation of the inverted-direction signal noted in the Predictive Results subsection: in this cohort, students with sparser EMA and sensor data were less, not more, likely to cross the moderate-depression cutoff. With only seven positive cases the magnitudes are fragile and should be treated as diagnostic rather than as evidence for a counter-hypothesis.

**Table S1.** Effective dimensionality of the 89-feature missingness matrix. *M*_eff_ (Cheverud–Nyholt) is the effective number of independent tests.

| Quantity | Value |
| --- | --- |
| Number of missingness features | 89 |
| PCs to capture 50% of variance | 2 |
| PCs to capture 80% of variance | 7 |
| PCs to capture 95% of variance | 16 |
| Effective dimensionality (Cheverud-Nyholt) | 5.5 |
| Components with eigenvalue > 1 (Kaiser) | 12 |

**Table S2.** Multivariable OLS regression of PHQ-9 outcomes on five a priori continuous-sensor missingness features (cont_miss_mean, cont_pct_zero_days, cont_longest_gap, cont_cov_slope, cont_late_pct_zero) and pre-term PHQ-9 (*n* = 38).

| Outcome | Predictor | $\beta$ | 95% CI | SE | P |
| --- | --- | --- | --- | --- | --- |
| Post-term PHQ-9 | Intercept | +0.56 | [−2.53, +3.65] | 1.52 | 0.713 |
| Post-term PHQ-9 | cont miss mean | +0.02 | [−13.04, +13.07] | 6.40 | 0.998 |
| Post-term PHQ-9 | cont pct zero days | −42.76 | [−170.09, +84.57] | 62.43 | 0.498 |
| Post-term PHQ-9 | cont longest gap | +0.30 | [−1.50, +2.10] | 0.88 | 0.739 |
| Post-term PHQ-9 | cont cov slope | −160.69 | [−630.00, +308.62] | 230.11 | 0.490 |
| Post-term PHQ-9 | cont late pct zero | +4.41 | [−16.05, +24.88] | 10.03 | 0.663 |
| Post-term PHQ-9 | pre | +0.91 | [+0.63, +1.20] | 0.14 | <0.001 |
| Δ PHQ-9 | Intercept | +0.56 | [−2.53, +3.65] | 1.52 | 0.713 |
| Δ PHQ-9 | cont miss mean | +0.02 | [−13.04, +13.07] | 6.40 | 0.998 |
| Δ PHQ-9 | cont pct zero days | −42.76 | [−170.09, +84.57] | 62.43 | 0.498 |
| Δ PHQ-9 | cont longest gap | +0.30 | [−1.50, +2.10] | 0.88 | 0.739 |
| Δ PHQ-9 | cont cov slope | −160.69 | [−630.00, +308.62] | 230.11 | 0.490 |
| Δ PHQ-9 | cont late pct zero | +4.41 | [−16.05, +24.88] | 10.03 | 0.663 |
| Δ PHQ-9 | pre | −0.09 | [−0.37, +0.20] | 0.14 | 0.546 |

**Table S3.** Principal-components analysis: PC scores correlated with PHQ-9 outcomes (top ten components). The smallest uncorrected *P* across all twenty PC–outcome tests is *P* = 0.013 (PC6 vs Δ PHQ-9), which does not survive Benjamini–Hochberg correction across the twenty tests (*q* ≈ 0.13).

| PC | Var explained (%) | r with post-PHQ-9 | P (post) | r with Δ-PHQ-9 | P (Δ) |
| --- | --- | --- | --- | --- | --- |
| 1 | 32.6 | +0.27 | 0.102 | −0.13 | 0.447 |
| 2 | 24.3 | −0.07 | 0.656 | +0.02 | 0.894 |
| 3 | 7.2 | +0.07 | 0.697 | +0.05 | 0.768 |
| 4 | 6.0 | −0.19 | 0.254 | +0.14 | 0.409 |
| 5 | 5.0 | −0.09 | 0.589 | −0.06 | 0.715 |
| 6 | 3.4 | −0.18 | 0.286 | −0.40 | 0.013 |
| 7 | 3.2 | +0.17 | 0.316 | −0.06 | 0.720 |
| 8 | 2.9 | −0.08 | 0.615 | −0.16 | 0.335 |
| 9 | 2.4 | +0.29 | 0.081 | +0.18 | 0.270 |
| 10 | 1.9 | −0.08 | 0.614 | −0.02 | 0.907 |

**Table S4.** Boston Marathon bombing sensitivity analysis. The bombing produced a real cohort-wide coverage drop but did not change the missingness–PHQ-9 association substantively.

| Analysis | Statistic | P | n | Notes |
| --- | --- | --- | --- | --- |
| 14-day pre vs post bombing cohort coverage | paired $t = 4.07$ | <0.001 | 38 | pre mean = 0.899; post mean = 0.855 |
| cont_miss_mean $\beta$ on post-PHQ-9, full window | $\beta = +0.97 \pm 4.56$, $R^2 = 0.631$ | 0.832 | 38 | — |
| cont_miss_mean $\beta$ on post-PHQ-9, excluding $\pm 14$d bombing window | $\beta = +0.82 \pm 3.52$, $R^2 = 0.631$ | 0.817 | 38 | — |

**Table S5.** Power analysis for the within-person daily analysis.

| Quantity | Value |
| --- | --- |
| Total person-day observations | 2,186 |
| Number of participants (clusters) | 49 |
| Mean observations per cluster | 44.6 |
| Within-person ICC of daily sensor coverage | 0.420 |
| Effective sample size after design effect | 113 |
| Minimum detectable $\vert r\vert $ (cluster-corrected, $\alpha = 0.05$, power = 0.80) | 0.261 |
| Minimum detectable $\vert r\vert $ (cluster-naive, for comparison only) | 0.060 |

**Table S6.** Monte Carlo power simulation for the omnibus joint *F*-test of nine continuous-stream missingness rates against post-term PHQ-9, adjusted for pre-term PHQ-9, at *n* = 38. Each row reports the empirical rejection rate at *α* = 0.05 across 1,000 simulated cohorts in which post-term PHQ-9 was generated from the empirically estimated pre-PHQ-9 model plus an isotropically directed missingness signal calibrated to the target incremental *R*^2^. Type-I error at Δ*R*^2^ = 0 is 0.045 (nominal 0.05); 80% power is reached at incremental *R*^2^ ≈ 0.17.

| Target $\Delta R^2$ | Power at $\alpha = 0.05$ | Simulations |
| --- | --- | --- |
| 0.0 | 0.045 | 1000 |
| 0.025 | 0.121 | 1000 |
| 0.05 | 0.221 | 1000 |
| 0.075 | 0.327 | 1000 |
| 0.1 | 0.458 | 1000 |
| 0.125 | 0.574 | 1000 |
| 0.15 | 0.7 | 1000 |
| 0.175 | 0.806 | 1000 |
| 0.2 | 0.867 | 1000 |
| 0.25 | 0.951 | 1000 |
| 0.3 | 0.993 | 1000 |
| 0.4 | 1.0 | 1000 |
| 0.5 | 1.0 | 1000 |

**Table S7.** Imputation sensitivity for the predictive sample. The published *n* = 32 Ridge (miss + pre) result *R*^2^ = −0.07 is compared with an *n* = 38 version under Bayesian-ridge multiple imputation (*m* = 20) of the structurally undefined feature `ema_daily_cov_cv.`The pre-PHQ-9-only baseline reaches LOO *R*^2^ = +0.579 on the *n* = 38 sample, essentially identical to the +0.59 reported on *n* = 32; multiple imputation does not reverse the qualitative conclusion that missingness features add no incremental signal.

| Analysis | Sample | LOO $R^2$ | Note |
| --- | --- | --- | --- |
| Pre PHQ-9 only | $n=38$ full | +0.579 | No imputation needed (no NaN in predictors). |
| Ridge (miss + pre) | $n=32$ complete-case (published in Table 3) | -0.070 | 6 participants dropped for undefined `ema_daily_cov_cv`. |
| Ridge (miss + pre) | $n=38$ with multiple imputation ($m=20$) | -0.012 | IterativeImputer + Bayesian-Ridge; mean across 20 imputations, $sd=0.062$. |

**Table S8.** Positive-control experiment for the LOO ridge pipeline. A synthetic biomarker calibrated to a target incremental *R*^2^ above the pre-PHQ-9 baseline was injected as a single column and the headline LOO ridge analysis was re-run for 50 random seeds at each target level. Observed mean incremental *R*^2^ recovered the injected signal to within ∼ 10% of target, confirming the pipeline is well-calibrated to detect signals at clinically interesting magnitudes (Δ*R*^2^ ≥ 0.10).

| Condition | Target $\Delta R^2$ (vs pre-PHQ-9) | Observed $\Delta R^2$ (vs pre-PHQ-9) | Observed $R^2$ | Note |
| --- | --- | --- | --- | --- |
| Real: pre-PHQ-9 alone | — (reference) | — (reference) | +0.585 | gain over cohort mean: +0.655 |
| Injected fake biomarker | +0.10 | +0.086 | +0.672 | mean across 50 seeds; SD($\Delta$)=0.050 |
| Injected fake biomarker | +0.20 | +0.191 | +0.776 | mean across 50 seeds; SD($\Delta$)=0.039 |
| Injected fake biomarker | +0.30 | +0.297 | +0.882 | mean across 50 seeds; SD($\Delta$)=0.018 |

**Table S9.** Feature-engineering ablation. The headline LOO ridge analysis was re-run under six alternative parameterizations of the missingness feature-set. No parameterization yielded miss-only LOO *R*^2^ *>* 0 or miss-plus-pre LOO *R*^2^ exceeding the pre-PHQ-9-only baseline (LOO *R*^2^ = +0.585) by more than 0.01. The closest competitor is the aggregate cohort-level-summary block (24 features = eight summary types × three stream families), where adding it to pre-PHQ-9 changes LOO *R*^2^ by only +0.006.

| Parameterization | n features | LOO $R^2$ (miss only) | LOO $R^2$ (miss + pre) | $\Delta$ vs pre only |
| --- | --- | --- | --- | --- |
| All 89 features (manuscript primary) | 89 | -1.233 | +0.168 | -0.418 |
| Continuous-stream features only | 25 | -0.544 | +0.390 | -0.195 |
| Phone-log features only | 21 | -0.449 | +0.505 | -0.080 |
| EMA features only | 43 | -1.607 | +0.072 | -0.513 |
| Aggregate cohort-level summaries | 24 | -0.210 | +0.592 | +0.006 |
| PCA top-5 rotation | 5 | -0.209 | +0.474 | -0.111 |

**Table S10.** Calibration of LOO classifiers for binary post-PHQ-9 ≥10 (*n* = 32, 7 positive cases). Quartile bins of LOO predicted probability vs observed event rate. The pre-PHQ-9 baseline is reasonably well-calibrated; the missingness-only L2 logistic is anti-calibrated (highest-predicted-probability bin contains zero events; lowest-predicted-probability bin contains 38% events), a direct visualisation of the inverted-direction signal.

| Model | Quartile bin | Mean predicted prob | Observed event rate | n in bin |
| --- | --- | --- | --- | --- |
| Pre PHQ-9 only | 1 (lowest) | 0.01 | 0.0 | 8 |
| Pre PHQ-9 only | 2 | 0.05 | 0.14 | 7 |
| Pre PHQ-9 only | 3 | 0.12 | 0.11 | 9 |
| Pre PHQ-9 only | 4 (highest) | 0.68 | 0.62 | 8 |
| L2 logistic, missingness only | 1 (lowest) | 0.01 | 0.38 | 8 |
| L2 logistic, missingness only | 2 | 0.04 | 0.25 | 8 |
| L2 logistic, missingness only | 3 | 0.26 | 0.25 | 8 |
| L2 logistic, missingness only | 4 (highest) | 0.75 | 0.0 | 8 |

**Fig. S1.**
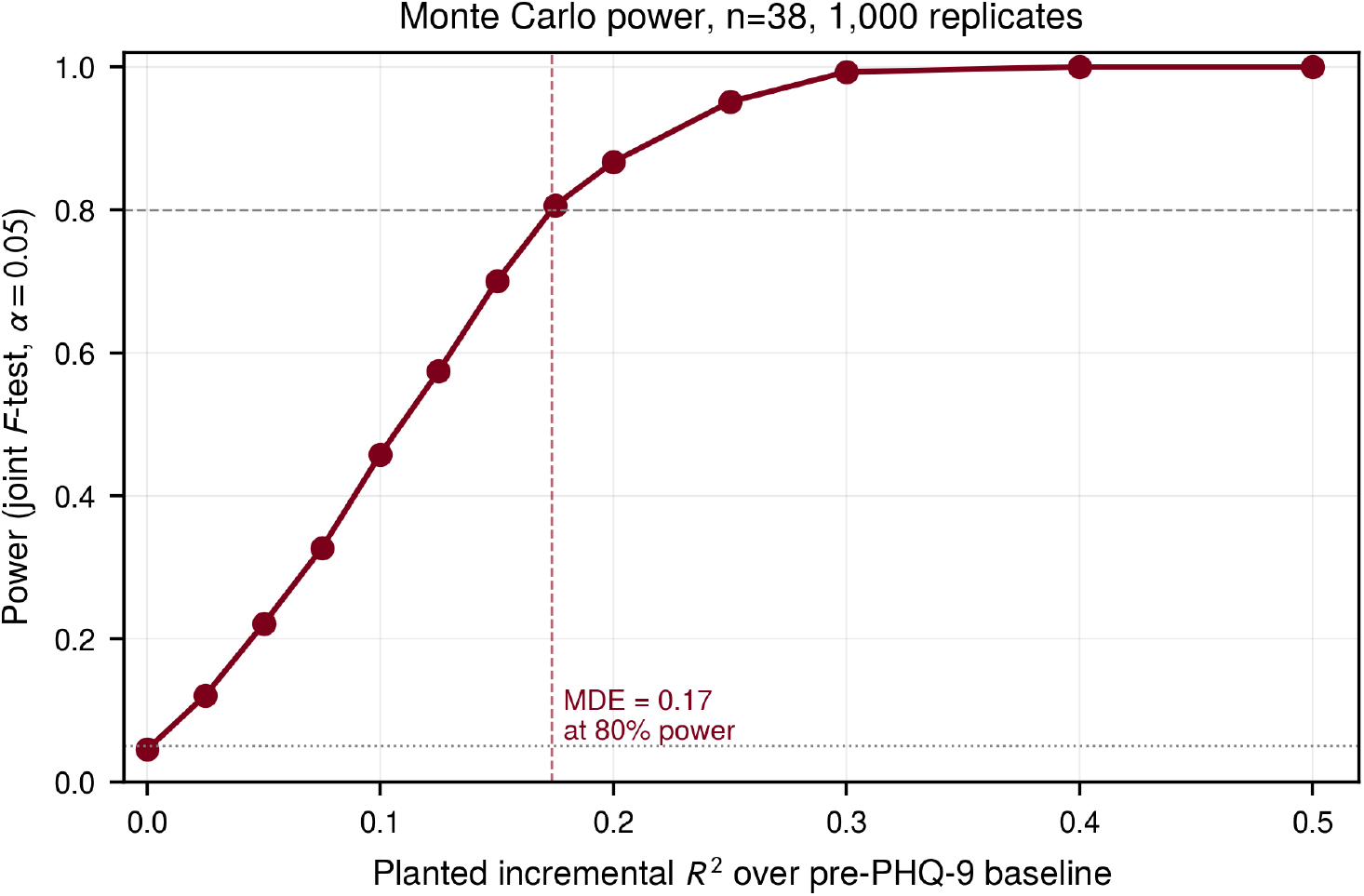
Monte Carlo power curve for the omnibus joint *F*-test of nine continuous-stream day-level missingness rates against post-term PHQ-9, adjusted for pre-term PHQ-9, at *n* = 38. Each point is the proportion of 1,000 simulated cohorts in which the joint *F*-test rejected at *α* = 0.05, where simulated post-PHQ-9 was generated from the empirically estimated pre-PHQ-9 OLS model plus an isotropically directed missingness signal calibrated to the target incremental *R*^2^. The empirical type-I error at Δ*R*^2^ = 0 was 0.045 (close to nominal). The 80% power crossing (dashed red vertical) is at incremental *R*^2^ = 0.17 above the pre-PHQ-9 baseline. *Takeaway:* the observed StudentLife null (*F* (9, 27) = 0.43, *P* = 0.91) is informative against missingness signals adding ≥17 percentage points of incremental out-of-sample variance, but cannot rule out small-to-moderate incremental signals.

**Fig. S2.**
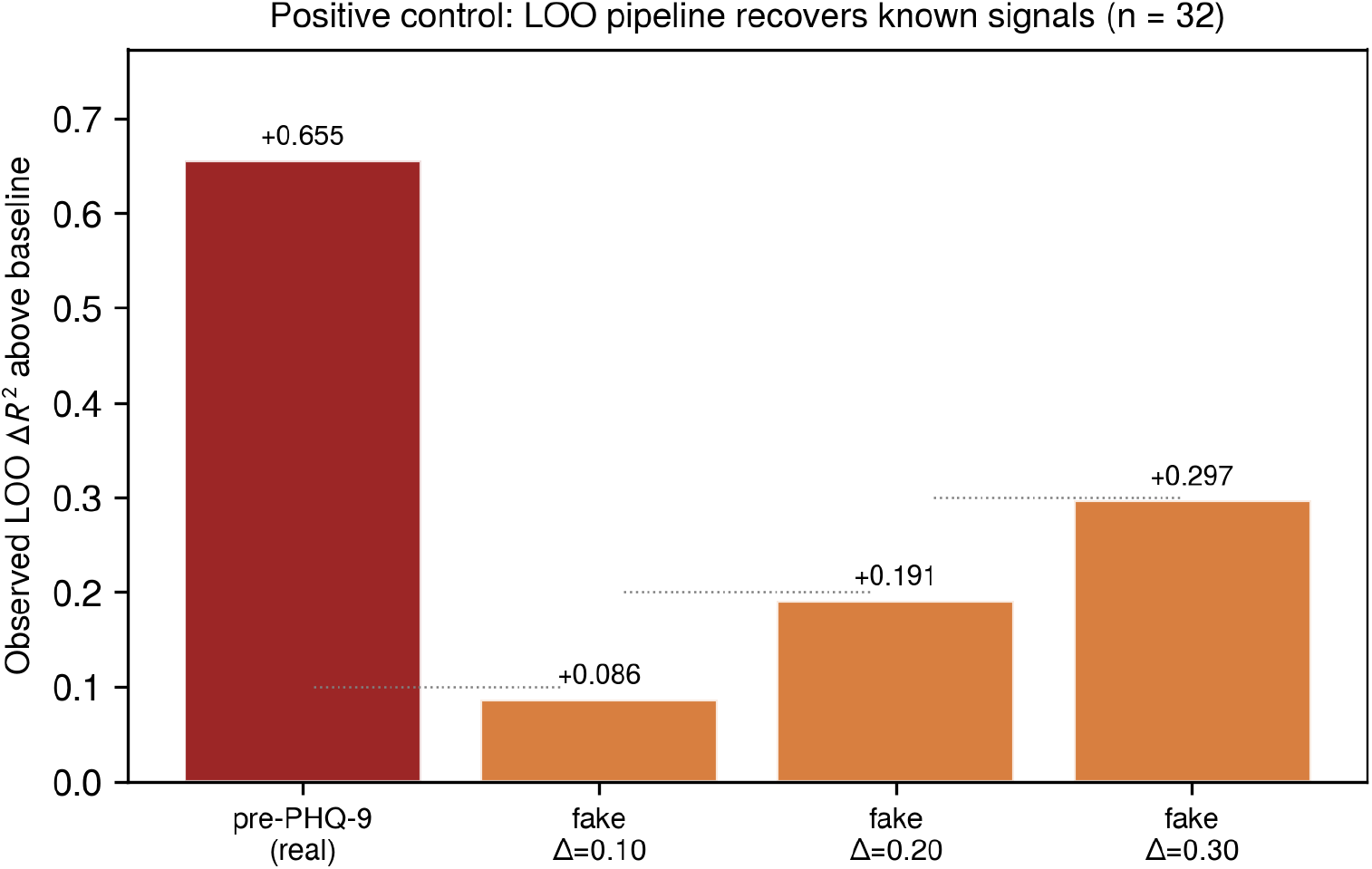
Positive-control experiment for the LOO ridge pipeline. The first bar shows the real LOO incremental *R*^2^ for pre-PHQ-9 over the cohort-mean baseline (+0.655). The remaining three bars show the LOO incremental *R*^2^ recovered when a synthetic biomarker calibrated to an injected target Δ*R*^2^ of 0.10, 0.20 or 0.30 is added as a single column to a pre-PHQ-9-only model (mean across 50 random seeds). Dotted grey lines mark the injection targets. *Takeaway:* the LOO pipeline recovers known signals to within ∼10% of target, so the observed null with the real missingness features is a property of the data and not a pipeline failure.

**Fig. S3.**
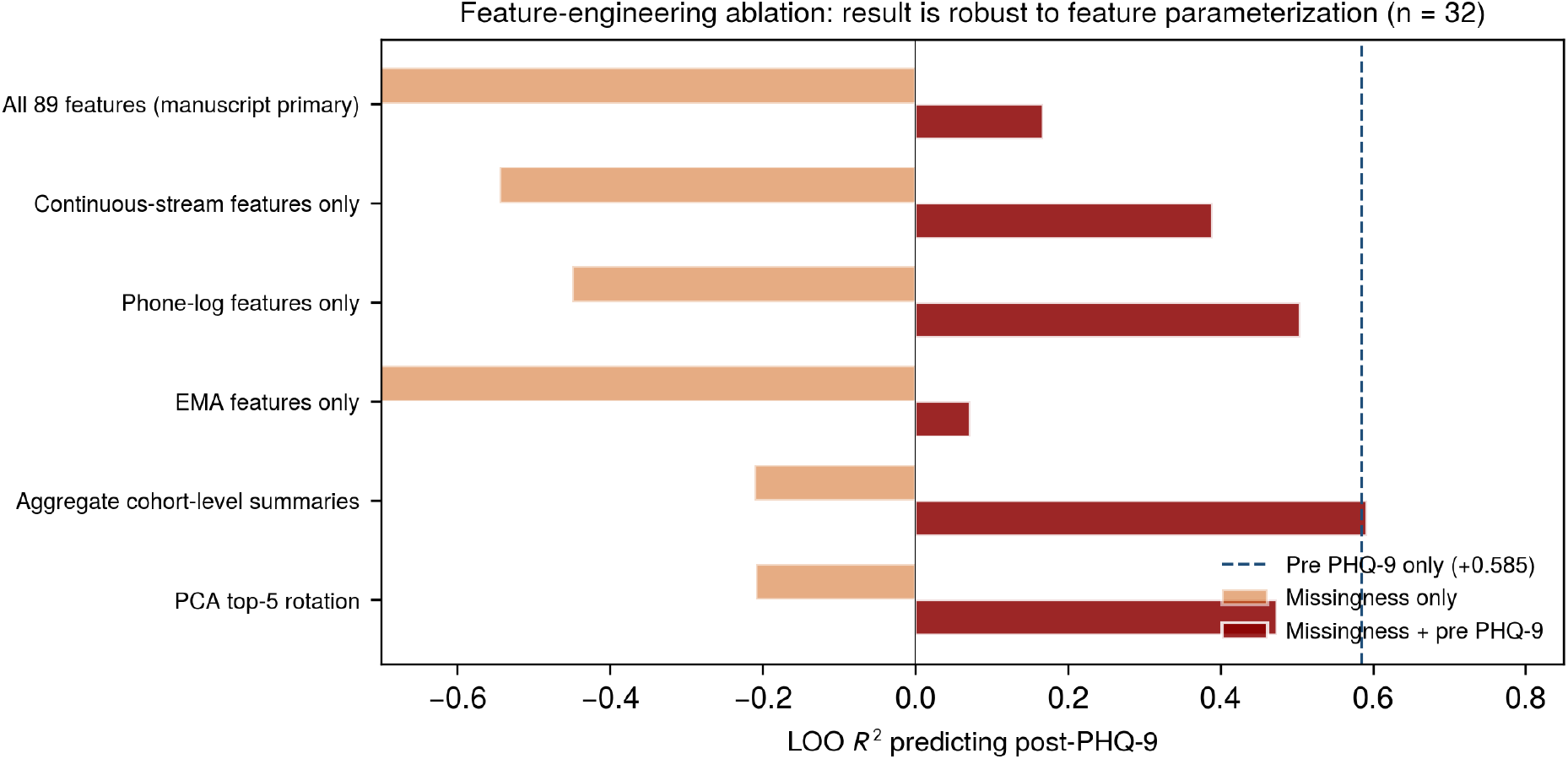
Feature-engineering ablation. LOO ridge *R*^2^ predicting post-term PHQ-9 from missingness alone (orange) and from missingness plus pre-PHQ-9 (red), under six alternative parameterizations of the missingness feature-set. The blue dashed line marks the pre-PHQ-9-only baseline (+0.585). *Takeaway:* no parameterization yields a missingness-only *R*^2^ above zero; no parameterization beats the pre-PHQ-9 baseline by more than 0.01 when both are combined. The result is robust to feature engineering.

**Fig. S4.**
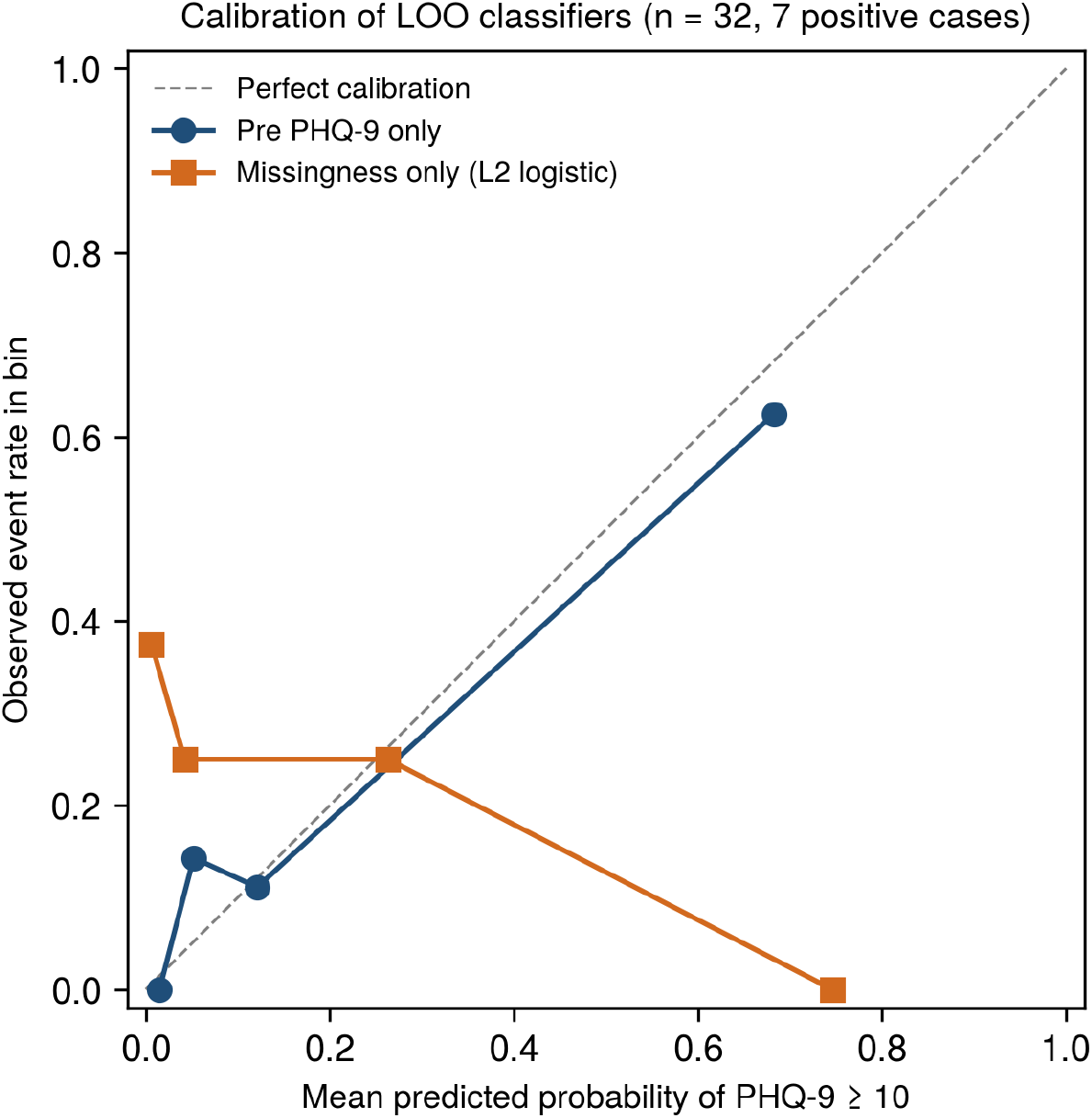
Calibration of LOO classifiers for binary post-PHQ-9 ≥10 (*n* = 32, 7 positive cases). Quartile bins of LOO predicted probability (x-axis) vs observed event rate (y-axis). The pre-PHQ-9 baseline (blue) hugs the diagonal of perfect calibration; the L2 logistic on missingness alone (orange) is anti-calibrated, with the highest-predicted-probability quartile containing zero observed events. *Takeaway:* this is a direct visualisation of the inverted-direction signal noted in the predictive Results subsection. With only seven positive cases, the magnitudes are fragile and should be treated as diagnostic of the inversion, not as evidence for a counter-hypothesis.

